# Does Subthalamic Deep Brain Stimulation Impact Asymmetry and Dyscoordination of Gait in Parkinson’s Disease?

**DOI:** 10.1101/2021.02.21.21251403

**Authors:** Deepak K Ravi, Christian R Baumann, Elena Bernasconi, Michelle Gwerder, Niklas König Ignasiak, Mechtild Uhl, Lennart Stieglitz, William R Taylor, Navrag B Singh

## Abstract

**Background:** Subthalamic deep brain stimulation is an effective treatment for selected Parkinson’s disease patients. Axial deficits including postural stability and gait characteristics are often altered after surgery, but quantitative gait-related therapeutic effects are poorly described.

**Objective:** The goal of this study was to systematically investigate modifications in asymmetry and dyscoordination of gait six-months post-operatively in patients with Parkinson’s disease, and compare the outcomes with preoperative baseline and to asymptomatic controls.

**Methods:** Thirty-two patients with Parkinson’s disease (19 with postural instability and gait disorder type, 13 with tremor-dominant disease) and 51 asymptomatic controls participated. Parkinson patients were tested prior to the surgery in both OFF and ON medication states, and six months post-operatively in the ON stimulation condition. Clinical outcome parameters and medication were compared to preoperative conditions. Asymmetry ratios, phase coordination index, and walking speed were assessed.

**Results:** Patients’ clinical outcomes as assessed by standard clinical parameters at six-months improved significantly, and levodopa-equivalent daily dosages were significantly decreased. STN-DBS increased step time asymmetry (hedges’ g effect sizes [confidence intervals] between pre- and post-surgery: 0.27 [-0.13,0.73]) and phase coordination index (0.29 [-0.08,0.67]). These effects were higher in the Postural Instability and Gait Disorder subgroup than the Tremor Dominant (step time asymmetry: 0.38 [-0.06,0.90] vs. 0.09 [-0.83,1.0] and phase coordination index: 0.39 [-0.04,0.84] vs. 0.13 [-0.76,0.96]).

**Conclusion:** This study provides objective evidence of how subthalamic deep brain stimulation increases asymmetry and dyscoordination of gait in patients with Parkinson’s disease, and suggests motor subtypes-associated differences in the treatment response.

## Introduction

Bipedal locomotion in humans, especially walking, is essential for living independently and high quality of life. In order to effectively walk, humans voluntarily activate neural centers propelling the musculoskeletal dynamics. There is surmounting evidence indicating that cortical centers are responsible for movement initiation, while ‘ongoing’ movements are steadily regulated via subcortical regions within the basal ganglia and the brain stem [1, 2]. These regions are responsible for providing internal cues to cortical and sub-cortical regions e.g. the pre-motor and supplementary motor area, accounting for symmetry and coordination between limbs, while allowing asymmetric control when needed (e.g. to negotiate obstacles during walking) [3]. Furthermore, symmetry and coordination between the upper and lower limbs are among the characteristic features required to generate balanced stepping, economical walking behaviour, but also maintaining speed. Interestingly, mild asymmetry in the walking patterns of asymptomatic healthy individuals is quite common and plausibly functional [4]. It is likely that such asymmetrical patterns during walking persist as a result of the natural hemispheric functional specificities in the relative contribution of lower limbs to propulsion (facilitate forward progression) and braking (facilitate postural stabilization) [4, 5]. In patients with movement disorders such as Parkinson’s disease (PD), asymmetry markedly increases and has been directly related to poor walking ability [6].

Parkinson’s disease is a progressive neurodegenerative disorder associated with neuronal loss and functional impairment of several regions of the central and autonomic nervous system. Traditionally, the degeneration of dopaminergic neurons in the substantia nigra pars compacta (part of basal ganglia, a subcortical area involved in locomotion [7]) are prominently involved in the cardinal motor deficiencies of the disease (bradykinesia/akinesia, tremor and rigidity). Apart from motor and non-motor symptoms, individuals with PD also experience substantial deficits with regards to bilateral function of walking, manifested by asymmetry ([8, 9]) and impaired coordination (or dyscoordination) during walking [8-10]. Importantly, individuals with these impairments also have a high tendency for adverse episodes such as freezing of gait (FOG) and falls [9, 11-13]. The underlying pathophysiology leading to these deficits are multifactorial and have not yet been completely understood. We may conceivably (albeit inconclusively) attribute these deficits to a bilateral but asymmetric neurodegenerative process that is characteristic of PD (asymmetrical motor function may persist over the years despite bilateral disease progression [14]). Patients with PD (PwPD) also appear to have reduced corpus callosum function, which interferes with the normal bilateral coordination of limb movements during walking [15]. Despite increased recognition of its prevalence, further research is clearly required to cement our understanding of asymmetrical presentation of walking dysfunctions in PwPD.

Deep brain stimulation (DBS) that targets the subthalamic nucleus (STN) of the basal ganglia has shown therapeutic potential as an adjunct to pharmacotherapy for alleviating motor symptoms of PD [16]. While the treatment appears effective in providing relief to the cardinal signs of the disease, the outcomes on gait remain inconclusive [17, 18]. Extensive evidence from long-term follow up studies has shown that certain aspects of gait function improve initially postoperatively, but then again progressively worsen [19]. Most of these studies have investigated improvement in gait function using Movement Disorder Society - Unified Parkinson’s Disease Rating Scale (MDS-UPDRS-III) clinical motor scores [20]. Although MDS-UPDRS-III is an internationally accepted and widely used clinical assessment scale (rapidly administered to measure clinically relevant outcomes), it only includes very few evaluations on walking ability or gait. The subtleties involved with controlling gait such as symmetry and coordination are important but remain unevaluated in standardized MDS-UPDRS-III assessments. Interestingly, such gait features could be used as ecologically valid biomarkers for both reliable candidate selection and assessing treatment endpoints thereafter. In this context, only a handful of studies - largely with acute ON vs OFF stimulation settings - have used instrumented and objective measures of gait (a review of studies is presented in [17, 18]). The analysis of such literature has shown consistently good effects on walking speed and stride length. However, varying reports on other gait parameters (e.g. cadence and stride time) and lack of long-term objective follow-ups preclude concrete conclusions with respect to therapeutic benefits of DBS.

In clinics, the practice of using bilateral DBS lead implantation is quite common, with surgical targets and stimulation settings often optimized to achieve best effects on upper limb tremor, rigidity etc., while minimizing relevant adverse effects (can be referred to as ‘clinically determined settings’). However, despite the prevalence of asymmetrical stimulation settings for bilateral DBS, functional asymmetry, particularly in the lower limbs, often persists postoperatively, and in some case, increases after surgery [21]. Importantly, insufficient attention to asymmetry may also be responsible for freezing of gait (FOG) and fall episodes in a subgroup of patients after the surgery [22, 23]. These deficiencies are possibly due to the obvious limitations regarding the lack of consideration of objective walking outcomes in clinically determined settings. As mobility and functional gait are critical for health and quality of life, an in-depth account of stimulation to gait and clinical outcome relations seems then essential for developing a comprehensive DBS therapy. As part of a larger study designed to investigate the effects of bilateral DBS on cardinal symptoms of PD, our project investigated gait performance as secondary outcomes of the treatment. In this paper, our primary objective was to systematically investigate alterations in asymmetry and dyscoordination of gait six-months post-DBS in PwPD. Our secondary objective was to examine whether the PD disease subtypes, including tremor dominant (TD), postural instability and gait disorder (PIGD), and indeterminate types show distinct therapeutic effects on gait and clinical outcomes.

## Materials and Methods

### Study design

Thirty-two PwPD (26 males, 6 females, with mean age 60.2 (SD 9.6) years, PD duration: 10.3 (5.0) years, preoperative MDS-UPDRS-III in OFF-Medication: 39.97 (12.51), Table 1 and Supp. Table 1) recruited at the University Hospital of Zurich, as well as 51 asymptomatic controls (22 males, 29 females, mean age 66.6 (10.7)) participated voluntarily in this study. The detailed surgical protocol and post-operative management of patients are provided in Supp. Methods 1. Stimulation parameters are presented in Supp. Table 2.

**Table 1:** Baseline and Longitudinal Demographics and Clinical Characteristics.

| Characteristic | Healthy Controls (N=51) | PwPD Pre DBS (N=32) | PwPD Post DBS (N=32) |
| --- | --- | --- | --- |
| Age (yr) | 66.6 (10.7) | 60.2 (9.6) | 60.9 (9.7) |
| Male/Female | 22/29 | 27/5 | 27/5 |
| Weight (kg) | 68.1 (12.2) | 77.0 (13.7) | 78.6 (11.7) |
| Height (cm) | 168.9 (8.8) | 176.0 (6.8) | 175.9 (6.7) |
| Age of onset of disease (yr) | - | 49.9 (9.3) | - |
| Disease duration (yr) | - | 10.3 (5.0) | - |
| H&Y stage | - | 2 (1 – 3) | 2 (1 – 2.5) |
| MDS-UPDRS based clinical subtypes | - | TD (N=13)<br>PIGD (N=19)<br>Indeterminate (N=0) | - |
| Symptom Side | - | Left-sided symptom onset (N=18)<br>Right-sided symptom onset (N=14) | - |
| Handedness | - | Right handedness (N=30)<br>Forced right handedness (N=2) | - |

Values expressed as mean (standard deviation) or median (lower limit – higher limit)
Abbreviations: MDS-UPDRS - Movement Disorder Society - Unified Parkinson's Disease Rating Scale; PwPD - Patients with Parkinson's disease; DBS - Deep Brain Stimulation; H&Y - Hoehn & Yahr; TD - Tremor Dominant; PiGD - Postural Instability and Gait Disorder

### Data collection

#### Clinical Assessment

Extrapyramidal motor symptoms were assessed using the MDS-UPDRS III, evaluated twice pre-surgery – in the ON and OFF medication conditions – as well as once around six months post-surgery in the medication and stimulation ON condition. MDS-UPDRS I, II and IV were also performed once (OFF medication condition) before surgery and once after surgery. In addition, dopaminergic treatment was recorded as levodopa equivalent daily dose. The clinical subtypes of PwPD such as TD, PIGD, and indeterminate types were also identified [24]. The predominant symptom side was identified by medical history and during the clinical examination and substantiated using the MDS-UPDRS asymmetry: difference between left and right motor scores of MDS-UPDRS Part III (items 3.3 - 3.8 and 3.15 - 3.17).

#### Objective Gait Measures

PwPD were tested prior to surgery in the medication ON state, and once around six months after the surgery in the ON medication and ON stimulation condition. Here, most subjects in the POST condition were still taking a clinically adapted dose of medication (Supp. Methods 1), except for three subjects that were completely OFF medication. All participants were instructed to walk (barefoot and self-selected speed) continuously for 10 minutes in an eight-shape around two marked spots 10 meters apart [25]. A three-dimensional motion capture camera system (10 cameras; 61 markers; 100 Hz; Vicon Nexus, version 2.3/2.8.2, Oxford Metrics, UK) was used to record the movements.

### Data analyses

#### Pre-processing

The raw kinematic data (from 2 × 7-meter straight sections of the “8-walk”) were low pass filtered (zero phase fourth order Butterworth with cut-off frequency of 25 Hz). The gait events (heel strikes and toe-offs) were automatically extracted using a custom algorithm based on foot velocity.

#### Asymmetry Ratio

Spatiotemporal gait measures of step length, step time, swing time and stance time (Supp. Methods 2) were evaluated for each foot separately. All outliers (defined as intervals outside ± 4 median absolute deviation (MAD) away from the median) were eliminated. For each subject, we then determined which foot had the larger vs. smaller amplitude (e.g. longer vs shorter mean step length) for evaluating the asymmetry ratios:

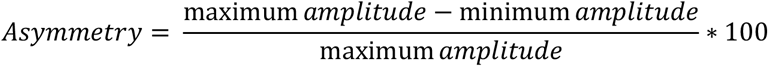

#### Phase Coordination Index

Bilateral dyscoordination in left-right stepping phase was analysed using the phase coordination index (PCI [26], Supp. Methods 2) Lower PCI values reflect a more accurate and consistent left-right stepping phase generation.

**Walking speed** was additionally calculated for all individuals.

### Statistical analyses

Clinical scores: Changes in the overall clinical scores were computed as: [(Pre-surgery scores - Post-surgery scores)/Pre-surgery scores] × 100%. The Wilcoxon signed-rank test was used to determine difference between the scores. Significance was set at *p* < 0.05.

Changes to individual items (and composite of sub-items) was computed as mean differences [confidence intervals]. The procedure for obtaining bootstrap confidence intervals [27] is detailed in Supp. Methods 3. These changes are not tested for significance because of large number of ties (zero differences) in the data.

Gait Characteristics: For the investigation of the neuromodulatory effects of DBS treatment on asymmetry and dyscoordination, effect sizes (Hedges’ g [27]) with confidence intervals (Supp. Methods 3) were calculated. The effect sizes (ES) were interpreted as: ES≤0.2 a negligible effect, 0.2 < ES ≤ 0.5 small to medium effect, 0.5 < ES ≤ 0.8 medium to large effect, and > 0.8 very large effect [28]. The rationale for including a control group is to demonstrate the importance of the direction of intervention effects (e.g. modification in asymmetry with respect to physiological asymmetry observed in asymptomatic controls). The *p* value of the two-sided permutation t-test is reported in case of significant differences (Supp. Methods 3, [27]). Significance was set at *p* < 0.05.

#### Symptom side and Gait Outcomes

Using a Sankey diagram, we visually represented the link between gait outcomes and the symptom side (e.g. does the leg with shorter step length always match with the symptom side at the beginning of the disease?) and the change in this relationship in response to the treatment.

All analyses were conducted in Matlab (v2019a, The Mathworks Inc., USA) and R (Version 1.2.5033, The R Foundation for Statistical Computing, Austria).

## Results

### Clinical characteristics

In the standardized evaluation of motor symptoms of the disease, MDS UPDRS-III score improved from a baseline Off Med value of 39.97 (12.5) by 58 percent at 6 months after surgery (Table 2). In comparison to the baseline Off Med, at 6 months the scores for UPDRS-I improved by 27 percent (range: −100 to 89), those for UPDRS-II improved by 46 percent (range: 0 to 90), and those for UPDRS-IV improved by 70 percent (0 to 100). All these functional improvements were significant (*p* < 0.001). Postoperatively, the levodopa-equivalent daily dosage also reduced significantly (*p* < 0.001) from 1118.8 (511.55) mg at baseline to 365.31 (219.86) mg at 6 months after surgery (Table 2).

**Table 2:** Effect of Bilateral Stimulation of the Subthalamic Nucleus on MDS-UPDRS scores.

| Characteristic | PwPD Pre DBS<br>Off Med | PwPD Pre DBS<br>On Med | PwPD Post DBS<br>On Med* | PwPD Pre DBS<br>Off Med vs<br>PwPD Post DBS<br>On Med* |  |  |  |  |
| --- | --- | --- | --- | --- | --- | --- | --- | --- |
| Overall Scores |  |  |  |  |  |  |  |  |
| MDS-UPDRS I | 10.13 (4.32) | - | 6.34 (2.70) | 26.86 (36.50)<sup>†</sup> |  |  |  |  |
| MDS-UPDRS II | 13.44 (4.65) | - | 7.38 (3.90) | 46.09 (23.10)<sup>†</sup> |  |  |  |  |
| MDS-UPDRS III | 39.97 (12.5) | 19.3 (7.4) | 16.5 (7.7) | 57.53 (15.68)<sup>†</sup> |  |  |  |  |
| MDS-UPDRS IV | 7 (3.30) | - | 1.81 (2.26) | 70.02 (32.23)<sup>†</sup> |  |  |  |  |
| Levodopa<br>equivalent daily<br>dose (mg/day) | - | 1118.8 (511.55) | 365.31 (219.86) | 64.50 (19.79)<sup>†</sup> |  |  |  |  |
| Subscores and Individual Items |  |  |  |  |  |  |  |  |
| Sub-items 3.15a<br>till 3.18, Sum of<br>Tremor (0-40) | 7.56 (6.32) | 2.06 (3.39) | 3.65 (3.50) | 3.91 [2.41, 5.47] |  |  |  |  |
| Sub-items 3.10<br>till 3.12, Sum of<br>PIGD (0-12) | 2.41 (2.23) | 1.06 (0.83) | 1.16 (1.50) | 1.19 [0.59, 2.38] |  |  |  |  |
| 2.10. Item<br>Tremor (0-4) | 1.19 (0.95) | - | 0.81 (0.74) | 0.38 [0.13, 0.59] |  |  |  |  |
| 2.12. Item<br>Walking and<br>Balance (0-4) | 1.13 (0.48) | - | 0.84 (0.63) | 0.28 [0.13, 0.41] |  |  |  |  |
| 2.13. Item<br>Freezing (0-4) | 0.66 (0.85) | - | 0.23 (0.49) | 0.44 [0.13, 0.72] |  |  |  |  |
| 3.10. Item Gait<br>(0-4) | 1.38 (0.70) | 0.78 (0.41) | 0.68 (0.53) | 0.66 [0.44, 0.97] |  |  |  |  |
| 3.11. Item<br>Freezing (0-4) | 0.41 (0.82) | 0.09 (0.29) | 0.23 (0.61) | 0.19 [-0.03, 0.56] |  |  |  |  |
| 3.12. Item<br>Postural Stability<br>(0-4) | 0.63 (0.96) | 0.19 (0.46) | 0.29 (0.73) | 0.34 [0.00, 0.72] |  |  |  |  |
| Clinical subtypes |  |  |  |  |  |  |  |  |
| Characteristic | PwPD Pre DBS<br>Off Med |  | PwPD Pre DBS<br>On Med |  | PwPD Post DBS<br>On Med* |  | PwPD Pre DBS<br>Off Med vs<br>PwPD Post DBS<br>On Med* |  |
|  | TD | PIGD | TD | PIGD | TD | PIGD | TD | PIGD |
| Sub-items 3.15a<br>till 3.18. Sum of<br>Tremor (0-40) | 13.69<br>(4.18) | 3.37<br>(3.47) | 3.54<br>(4.53) | 1.05<br>(1.67) | 6.38<br>(3.43) | 1.67<br>(1.80) | 7.31 [5.62,<br>9.54] | 1.58 [0.16,<br>3.16] |
| Sub-items 3.10<br>till 3.12. Sum of<br>PIGD (0-12) | 1.23<br>(0.58) | 3.21<br>(2.57) | 0.77<br>(0.42) | 1.26<br>(0.96) | 0.77<br>(0.89) | 1.50<br>(1.77) | 0.46 [-0.31,<br>0.77] | 1.68 [0.79,<br>3.58] |

Values expressed as mean (standard deviation)
\*3 patients were in OFF medication state
Change on MDS-UPDRS I to IV and Levodopa equivalent daily dose are provided as mean (standard deviation) percentages. <sup>†</sup> $p < 0.001$ ; The $p$ values reported denotes level of statistical significant difference
Change on subscores, individual items, clinical subtypes are provided as mean differences [confidence intervals].
Abbreviations: MDS-UPDRS - Movement Disorder Society - Unified Parkinson's Disease Rating Scale; PwPD - Patients with Parkinson's disease; DBS - Deep Brain Stimulation; TD - Tremor Dominant; PIGD - Postural Instability and Gait Disorder

A subgroup analysis was performed on MDS-UPDRS III items that address clinical subtypes [29]. The average **MDS**-**UPDRS TD/PIGD ratio** for TD group was 3.28 (1.34) and for PIGD group 0.33 (0.29). The mean difference [95% confidence intervals] between the groups was 2.95 [2.32, 3.79]. The composite scores for tremor improved by 3.91 [2.41, 5.47], and those for posture and gait improved by 1.19 [0.59, 2.38], both near to 50% group-level improvement. **MDS-UPDRS Asymmetry:** The difference between left and right MDS-UPDRS III motor subscores was 2.29 (7.59) in PwPD, 4.23 (8.69) in TD group and 0.89 (6.32) in PIGD group.

### Gait characteristics PwPD

PwPD had higher asymmetry ratios (highest effect size reported for step length asymmetry, ES [CI]: 0.56 [0.06, 1.03], *p* < 0.05) and higher PCI (0.81 [0.3, 1.26], *p* < 0.05) before surgery compared to the asymptomatic control group (Supp. Table 3).

Increase in step time asymmetry (0.27 [-0.13, 0.73]) and PCI (0.29 [-0.08, 0.67]) was observed post-surgery relative to pre-surgical status in PwPD. Differences in all other gait characteristics between pre- and post-surgery were negligible (Table 3).

**Table 3:** Effect of Bilateral Stimulation of the Subthalamic Nucleus on Gait Characteristics.

| Characteristic | PwPD Pre DBS | PwPD Post DBS | Healthy Controls | PwPD Pre DBS vs PwPD Post DBS |
| --- | --- | --- | --- | --- |
| **Asymmetry (%)** |  |  |  |  |
| Step length | 4.81 (4.19) | 4.60 (3.50) | 2.90 (2.69) | 0.05 [-0.38,0.32] |
| Step time | 2.79 (2.32) | 3.48 (2.61) | 1.93 (1.70) | 0.27 [-0.13,0.73] |
| Swing time | 3.35 (4.19) | 3.76 (4.07) | 2.36 (1.92) | 0.10 [-0.24,0.49] |
| Stance time | 2.06 (2.66) | 2.36 (2.72) | 1.48 (1.20) | 0.11 [-0.21,0.51] |
| **Dyscoordination (%)** |  |  |  |  |
| PCI | 3.06 (1.06) | 3.42 (1.41) | 2.29 (0.82) | 0.29 [-0.08,0.67] |
| **Other gait measures** |  |  |  |  |
| Walking Speed(cm/s) | 117.09 (18.21) | 119.11 (18.42) | 120.95 (14.31) | 0.11 [-0.11,0.36] |

Values expressed as mean (standard deviation)
Changes are provided as hedges' g effect sizes [confidence intervals].
Abbreviations: PCI - Phase Coordination Index; PwPD - Patients with Parkinson's disease; DBS - Deep Brain Stimulation;
Please refer to Supp. Table 3 for comparison scores (effect sizes) against healthy controls.

### Gait characteristics Clinical Subtypes

#### Between Subtypes

The magnitude of difference between the TD and PIGD subtypes were larger for PCI and walking speed (both ES’s > 0.50) before surgery. The differences between the subtypes increased post-surgery relative to pre-surgical status in all the gait characteristics (notably step time asymmetry 0.40 [-0.33, 0.972] to 0.69 [-0.03, 1.33] and PCI 0.60 [-0.10, 1.13] to 0.78 [0.06, 1.35], *p* < 0.05) except walking speed (−0.55 [-1.27, 0.22] to −0.32 [-0.98, 0.40]), Table 4.

**Table 4:** Effect of Bilateral Stimulation of the Subthalamic Nucleus on Gait Characteristics: Clinical Subtypes.

| Characteristic | PwPD Pre DBS |  | PwPD Post DBS |  | Between Subtypes |  | Within Subtypes |  |
| --- | --- | --- | --- | --- | --- | --- | --- | --- |
|  | TD | PIGD | TD | PIGD | TD Pre vs PIGD Pre | TD Post vs PIGD Post | TD Pre vs TD Post | PIGD Pre vs Post |
| **Asymmetry (%)** |  |  |  |  |  |  |  |  |
| Step length | 4.52 (4.64) | 5.00 (3.85) | 4.24 (2.17) | 4.85 (4.15) | 0.11 [-0.74, 0.80] | 0.17 [-0.51, 0.75] | 0.07 [-0.60,0.65] | 0.04 [-0.46,0.39] |
| Step time | 2.22 (1.84) | 3.18 (2.52) | 2.41 (2.06) | 4.21 (2.70) | 0.40 [-0.33, 0.972] | 0.69 [-0.03, 1.33] | 0.09 [-0.83,1.0] | 0.38 [-0.06,0.90] |
| Swing time | 2.81 (3.85) | 3.71 (4.37) | 3.05 (2.85) | 4.25 (4.66) | 0.20 [-0.68, 0.75] | 0.28 [-0.45, 0.80] | 0.06 [-0.57,0.67] | 0.11 [-0.29,0.67] |
| Stance time | 1.73 (2.23) | 2.28 (2.90) | 1.70 (1.46) | 2.82 (3.24) | 0.20 [-0.63, 0.72] | 0.40 [-0.28, 0.84] | 0.01 [-0.64,0.59] | 0.17 [-0.22,0.69] |
| **Dyscoordination (%)** |  |  |  |  |  |  |  |  |
| PCI | 2.67 (0.73) | 3.32 (1.16) | 2.79 (0.90) | 3.86 (1.52) | 0.60 [-0.10, 1.13] | **0.78 [0.06, 1.35] \*** | 0.13 [-0.76,0.96] | 0.39 [-0.04,0.84] |
| **Other gait measures** |  |  |  |  |  |  |  |  |
| Walking Speed(cm/s) | 123.14 (15.59) | 112.96 (18.71) | 122.77 (15.52) | 116.60 (19.78) | -0.55 [-1.27, 0.22] | -0.32 [-0.98, 0.40] | -0.02 [-0.34,0.34] | 0.18 [-0.11,0.54] |

Values expressed as mean (standard deviation)
Changes are provided as hedges' g effect sizes [confidence intervals].
\* $p < 0.05$ ; The $p$ values of the two-sided permutation t-test
Abbreviations: PCI - Phase Coordination Index; PwPD - Patients with Parkinson's disease; DBS - Deep Brain Stimulation; TD - Tremor Dominant; PIGD - Postural Instability and Gait Disorder

#### Within Subtypes

Step time asymmetry (0.38 [-0.06, 0.90]) and PCI (0.39 [-0.04, 0.84]) increased in the PIGD group following surgery. Differences in all other gait characteristics within subtypes, between pre- and post-surgery are negligible (Table 4).

**In comparison to healthy controls**, the PIGD group registered significant differences (*p* < 0.05) in the following gait characteristics: step length asymmetry (−0.67 [-1.35, −0.06]), step time asymmetry (−0.62 [-1.23, −0.03]) and PCI (−1.08 [-1.64, −0.42]) before surgery; step length asymmetry (−0.60 [-1.3, 0.01]), step time asymmetry (−1.1 [-1.8, −0.42]), swing time asymmetry (−0.63 [-1.2, −0.03]), stance time asymmetry (−0.66 [-1.22, −0.03]) and PCI (−1.45 [-2.14, −0.75]) after surgery, Supp. Table 3.

#### Symptom side

The symptom side diagnosed through medical history matched with the MDS-UPDRS based predominant symptom side in 84.3% (27 out of 32 patients). Interestingly, all the 5 mismatched patients belonged to the PIGD group. Based on the MDS-UPDRS, 56.3% (18 out of 32 patients) experienced their first symptom on the left side of the body and were still more markedly affected on this side. As normally expected, the predominant symptom side matched with the side having smaller amplitude gait parameters: in 16 out of 32 (for step length), but only in 14 out of 32 (for step time) and in 8 out of 32 (for swing time). After surgery, the previously predominant symptom side reversed: in 10 out of 32 (for both step length and step time) and in 9 out of 32 (for swing time) PwPD patients (Figure 1).

**Figure 1:**
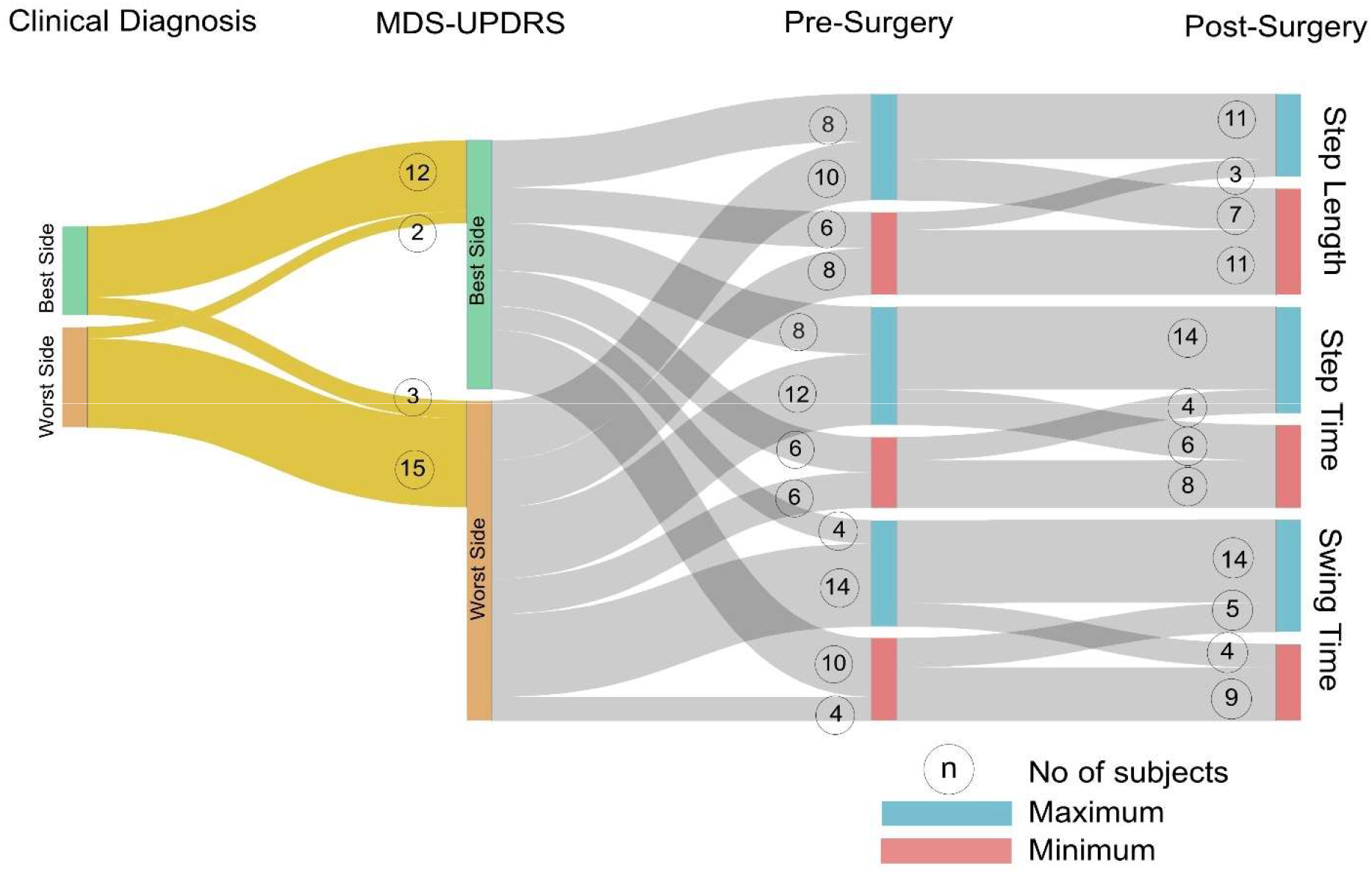
Association between Gait Characteristics and Predominant Symptom Side. Note: The flows marked in yellow are provided as an illustrative example and should be interpreted as follows: The symptom side clinically diagnosed during the first observation by the study neurologist matched with the MDS-UPDRS based symptom side in 27 patients. Another example interpretation: Before surgery, the leg with longer step length (‘Maximum’) matched with the non-dominant symptom side (‘Best Side) in 8 patients and with the predominant symptom side (‘Worst Side’) in 10 patients. Ideally, one would expect the leg with longer step length always match with the non-dominant symptom side of PD.

## Discussion

Outside of standard clinical outcome measures, it remains unclear how STN-DBS impacts objective and comprehensive motor characteristics on a long-term. Towards this understanding, our study reports a six-month follow-up of motor and gait outcomes after STN-DBS in PwPD. Our results revealed the following: 1) As anticipated, PwPD showed significant motor and non-motor benefits (MDS-UPDRS I through IV) after surgery, together with a significant reduction in dopaminergic medication. 2) STN-DBS induced increase in step time asymmetry and dyscoordination at follow up. 3) Group differences in baseline (before surgery) gait characteristics are critical to discretise treatment planning, as indicated by higher level of asymmetry, dyscoordination and slower walking speed in PIGD patients compared to the TD group. Six-month increase in step time asymmetry and dyscoordination after surgery were higher in the PIGD group than the TD group. Collectively, our study documents novel aspects of functional differences in DBS treatment responses guided by different motor subtypes with an eye toward their relevance as objective biomarkers for use in clinical settings.

Causal mechanisms behind motor asymmetry and side predominance in PwPD remain unsettled. Whilst most studies have assumed a main effect of natural differences in the numbers of nigral dopaminergic neurons, motor asymmetry have also been linked to handedness [30]. Similar to clinical motor symptoms, spatio-temporal parameters of gait in PwPD have also been found to be presented with asymmetry, over and above the physiological levels commonly reported in healthy individuals [4]; and independent of MDS-UPDRS asymmetry [11, 15]. Presumably, the control of these two functions are mediated by different anatomical or functional pathways in humans [31]. Asymmetry and bilateral dyscoordination during walking have traditionally been regarded as pathological in PwPD. Asymmetrical gait characteristics are often the first motor symptoms seen in these populations [32] and demonstrated as a clinical marker of prodromal PD [33]. Although, there is only a marginal agreement on the underpinnings of asymmetrical motor and gait symptoms in PwPD, such features are starting to provide essential movement biomarkers to guide diagnosis and treatment.

The primary motor symptoms of PD are highly responsive to dopaminergic replacement therapies. Nevertheless, the benefit to PwPDs, promoted by the administration of such therapies, tends to decline with the natural course of the disease, as wearing-off, dyskinesia and other forms of motor fluctuations occur and deteriorate [34]. STN-DBS is an effective and well-established treatment in ameliorating the cardinal symptoms in PwPD, as well as levodopa-induced motor complications, but also in allowing for significantly reducing dopaminergic drug intake. Similar to previous studies, we found that STN stimulation produced significant motor improvements as assessed by the MDS-UPDRS scores [16]. Currently available evidence suggests that DBS improves axial symptoms of PD only moderately, if ever, such as deficits in gait and postural control. STN is a complex subcortical motor network and regulate a myriad of motor features required in daily life of PwPD. Our current approaches to STN-DBS therapy are based on an inadequate assessment of such features; thus, hinder the making of any definitive recommendations about the therapy. In order to improve this state-of-knowledge in a systematic manner, our study investigated asymmetry and dyscoordination in gait of PwPD, both pre-operatively and 6-months post-surgery and compared these metrics to physiological levels assessed from a sample of asymptomatic controls.

DBS treatment seems like an ideal approach for improving asymmetry and dyscoordination of walking as it provides the opportunity to modify stimulation parameters for each hemisphere independently. On the other hand, we might expect that some degree of asymmetry may persist because of the apparent role of hemispheric dominance in the regulation of gait, but also asymmetrical stimulation settings in relation to symptom-side predominance [23, 35]. In contrast to these assumptions and despite the overall favourable outcome of DBS in this PwPD cohort, we observed that STN-DBS increased gait asymmetry and dyscoordination, when comparing pre versus post-surgery. Similar to our findings, one recent study [21] also reported increased gait asymmetry six-month post DBS compared to preoperative assessment. These results differ from a previous work that demonstrated a significant improvement in gait asymmetry in response to STN-DBS treatment [13]. The difference in results can be attributed primarily to the study design (Johnsen and colleagues only presented acute postoperative DBS ON vs OFF gait outcomes in OFF medication state). Postoperative microlesion effects (e.g. some permanent lesion related to electrode insertion) may introduce bias [36] and does not allow us to equate and compare the Pre-DBS state (of our study) to Off DBS state (in Johnsen et al. 2009). The differences can also be due to the definition of gait asymmetry (Johnsen and colleagues looked at spatial distance between heel to projected center of mass [13]). The increase in asymmetry and dyscoordination that we are reporting are calculated based on temporal metrics (step time asymmetry and PCI).

Gait impairments in PwPD, from the symptom onset to their advanced stages, including response to treatments, demonstrate patient as well as group (TD vs PIGD) specific traits [37]. In our study, the TD group had comparatively less impaired gait to the overall group of patients both before and after surgery. On the other hand, PIGD patients demonstrated a more pronounced increase (more than the longitudinal changes due to disease progression reported previously in PwPD [38]) in asymmetry and dyscoordination compared to the TD group due to DBS, consistent with a previous clinical investigation [37]. Differences in natural prognosis (for example, PIGD may naturally progress more quickly in disease course) likely underlie differences in treatment outcomes but also mechanisms among PwPD subtypes. Surprisingly, step length asymmetry did not show any group-specific differences in the gait outcomes. Interestingly, in our study, step time asymmetry and dyscoordination increased, however, step length asymmetry did not. This difference plausibly indicates a differential effect of STN-DBS on the temporal deficit or a possible adaptation of the temporal characteristics to allow a regulation of spatial characteristics: in this case step length asymmetry or possibly to optimize energy cost. In light of the accumulating evidence that these parameters may reflect distinct regulation but also mutual interactions, the hypotheses of a differential effect vs potential adaptation among these spatio-temporal characteristics needs further exploration [39].

One key limitation of our study is the lack of information about fall history of the participants. This limits the ability of our study to validate the outcomes directly with the fall status of the participants. Here, a previous study has reported a significant inverse relationship between PCI and the UPDRS item describing postural stability [10]. There were also limitations in the design of the study (gait being a secondary outcome) that did not allow us to replicate novel unilateral stimulation protocols suggested towards improving gait asymmetry and dyscoordination in patients with Parkinson’s disease [23]. Our findings show the efficacy of stimulation of the subthalamic nucleus in reducing motor symptoms and related clinical outcomes in PwPD at 6 months after surgery. Despite clinical improvement, the DBS treatment affected the asymmetry and dyscoordination of walking. In particular, PIGD patients demonstrated a more pronounced decline in these gait characteristics compared to the TD group due to DBS. Thus, we may hypothesize that clinical subtypes of PD underlie such treatment responses, thus providing relevant and interpretable insights into the mechanisms possibly regulating these subtle gait characteristics.

## Data Availability

Please contact the corresponding author for requests regarding data sharing.

## Acknowledgements

The authors would like to thank all the subjects for participating in the study and their families for the support provided.

## Author contributions

NKI, NBS, CRB and WRT conceived and designed the study. CRB, LS, MU coordinated the recruitment, pre-surgery and post-surgery of the participants. DKR, MG, EB and NKI coordinated the gait measurements. DKR, MG and EB analysed the data. DKR, NBS and WRT drafted the manuscript. CRB provided clinical content expertise while drafting and revising the manuscript. LS provided critical opinion and revision of the manuscript as a subject expert. All the authors reviewed and approved the manuscript for submission. WRT is the guarantor.

## Ethics approval

The study was approved (approval no: 2015-00141) by Zurich Cantonal Ethics Commission and carried out in accordance with the Declaration of Helsinki, the guidelines of Good Clinical Practice, and the Swiss regulatory authority’s requirements. The subjects all provided written, informed consent prior to participation.

## Supp. Methods 1: Surgical and post-operative procedures

Preoperative MR images of the entire cortex were obtained on a Philips Achieva 3T Scanner (Philips, AMS, Netherlands), and scans were fused on the Medtronic S7 surgical planning station (Medtronic plc, MN, USA). Thereafter, the volumetric image data was reformatted according to anterior and posterior commissures (AC-CP). Indirect target planning was initiated at X_lat_ = ±12 mm, Y_ap_ = −3 mm, Z_vert_ = −4 mm relative to the mid-commisural point (MCP), and then directly adapted to the hypo-intense zones associated with the nucleus ruber, the substantia nigra and the STN in the MR images of each individual PwPD to target the postero-dorso-lateral motoric portion of the STN. Intra-operatively, DBS lead penetration into the STN was guided by a stereotactic frame (Riechert-Mundinger, Inomed Medizintechnik GmbH, Germany). Target accuracy was perioperatively confirmed based on single-to multi-electrode micro-recording. Then, during test stimulation through a macro-electrode, we performed neurological testing of rigidity, tremor, and akinesia, while documenting the side effects. On this basis, the optimal sites for stimulation were determined before the neurosurgeon implanted the definitive stimulation leads (Medtronic Model 3389). After bilateral lead implantation in the STN, perioperative CT images were obtained to enable target reconstruction. The CT images were fused with the preoperative MRI scans, and the localization of the centers of all four lead contacts were reconstructed. A few days after implantation, stimulation was gradually started at the optimal contact locations determined during intraoperative neurological testing. In some patients, the stimulation site needed to be adapted to optimize clinical impact and to reduce the side effects. The stimulation was then optimized during a 2-4-week in-patient rehabilitation and over multiple out-patient follow-up examinations. In each patient, an optimal combination of electrical stimulation and reduced pharmacological medication was targeted to alleviate the cardinal symptoms of PD and their fluctuations until stable outcomes were reached (‘clinically determined settings’). Stimulation parameters at six months post-surgery are presented in Supp. Table 2.

## Supp. Methods 2: Definition of gait parameters

### Step length

is the anterior-posterior distance measured from the heel contact of one foot to the heel contact of the opposite foot.

### Step time

is the time elapsed from the heel contact of one foot to the heel contact of the opposite foot.

### Swing time

The swing phase is the non-weight-bearing portion of each gait cycle begins when the foot (toe) first leaves the ground and ends when the same foot (heel) touches the ground again.

### Stance time

The stance phase is the weight-bearing portion of each gait cycle begins when the foot (heel) first touches the ground and ends when the same foot (toe) leaves the ground. Stance time is the amount of time that passes during the stance phase of one leg.

### PCI

The PCI, expressed in percentage, is the sum of the coefficient of variation of phase and the mean absolute difference between the phase and 180°, normalized to 180°. Phase is the relative timing between the contra-lateral heel contacts.

### Walking speed

Walking speed is calculated from the distance covered by the sacrum marker between two consecutive gait cycles and the time taken.

## Supp. Methods 3: Bootstrap Confidence Intervals and Permutation t-test

### Bootstrap Confidence Intervals

We calculate the 95% Confidence Intervals of the mean difference by performing bootstrap resampling. We create multiple resamples (with replacement) from our set of observations and computes the effect size on each of these resamples. The bootstrap resamples of the effect size is used then to determine the 95% Confidence Intervals. Here we use 5000 resamples.

More Information: https://www.estimationstats.com/#/background

### Permutation t-test

For example, we measure a characteristic X for each individual from two groups P and C whose sample means are M_P_ and M_C_ and that we want to know whether X_P_ and X_C_ come from the same distribution. The permutation test is intended to verify whether the observed difference between the sample means M_P_ and M_C_ is large enough to reject, at some significance level, the null hypothesis that the data drawn from P is from the same distribution as the data drawn from C.

More Information: https://en.wikipedia.org/wiki/Resampling_(statistics)#Permutation_tests

**Supp. Table 1:** Demographics and Clinical Characteristics by Parkinson’s Disease Clinical Subtypes.

| **Characteristic** | **TD**<br>**(N=13)** | **PIGD**<br>**(N=19)** |
| --- | --- | --- |
| **Age (yr)** | 60 (10.4) | 60.4 (9.0) |
| **Male/Female** | 11/2 | 16/3 |
| **Weight (kg)** | 81.8 (11.3) | 73.7 (14.1) |
| **Height (cm)** | 176.2 (6.3) | 175.9 (7.1) |
| **Age of onset of disease (yr)** | 49.8 (10.2) | 49.9 (8.6) |
| **Disease duration (yr)** | 10.2 (6.1) | 10.4 (4.0) |

Values expressed as mean (standard deviation)
Abbreviations: TD - Tremor Dominant; PIGD - Postural Instability and Gait Disorder

**Supp. Table 2:** Stimulation Parameters at 6 months Follow-Up.

| Patient ID | Frequency [Hz] | Voltage [V] | | Contacts | | Pulse width [μs] | |
| --- | --- | --- | --- | --- | --- | --- | --- |
|  |  | left | right | left | right | left | right |
| P001 | 130 | 1.4 | 1.6 | 2-C+ | 10-C+ | 60 | 60 |
| P002 | 130 | 4.4 | 2.7 | 2-3+ | 10-C+ | 60 | 60 |
| P003 | 130 | 1.3 | 2 | 2-C+ | 10-C+ | 60 | 60 |
| P004 | 130 | 2.4 | 2.5 | 1-C+ | 9-C+ | 60 | 60 |
| P005 | 130 | 2.5 | 2.3 | 2-C+ | 10-C+ | 60 | 60 |
| P006 | 130 | 1.5 | 1.5 | 2-C+ | 10-C+ | 60 | 60 |
| P007 | 130 | 3.2 | 4.5 | 2-1+ | 10-C+ | 60 | 60 |
| P009 | 130 | 1.4 | 1.3 | 2-C+ | 10-C+ | 60 | 60 |
| P010 | 130 | 4.1 | 1.8 | 3-1+ | 10-C+ | 60 | 60 |
| P011 | 130 | 1.6 | 1.5 | 2-C+ | 10-C+ | 60 | 60 |
| P012 | 130 | 2.9 | 2.5 | 2-C+ | 10-C+ | 60 | 60 |
| P013 | 130 | 1.1 | 1.5 | 2-C+ | 10-C+ | 60 | 60 |
| P014 | 130 | 1.5 | 1.6 | 2-C+ | 10-C+ | 60 | 60 |
| P015 | 130 | 1.7 | 2.9 | 2-C+ | 10-C+ | 60 | 60 |
| P016 | 130 | 2 | 2 | 2-C+ | 10-C+ | 60 | 60 |
| P017 | 130 | 3.6 | 3.8 | 2-C+ | 11-C+ | 60 | 60 |
| P019 | 180 | 3.7 | 3.5 | 1-3+ | 10-9+ | 60 | 60 |
| P020 | 130 | 2.1 | 2 | 1-C+ | 10-C+ | 60 | 60 |
| P021 | 130 | 1.7 | 2 | 1-C+ | 9-10+ | 60 | 60 |
| P023 | 130 | 1.3 | 2.8 | 2-C+ | 10-C+ | 60 | 60 |
| P024 | 130 | 2.5 | 1.7 | 2-C+ | 10-C+ | 60 | 60 |
| P025 | 130 | 1.5 | 1.6 | 2-C+ | 10-C+ | 60 | 60 |
| P026 | 160 | 2 | 2 | 3-1+ | 11-9+ | 60 | 60 |
| P027 | 130 | 2.1 | 3.6 | 1-0+ | 10-9+ | 60 | 60 |
| P029 | 130 | 3.1 | 2.4 | 2-C+ | 10-C+ | 60 | 60 |
| P030 | 130 | 3.1 | 2.4 | 2-C+ | 10-C+ | 60 | 60 |
| P031 | 130 | 2.4 | 1.9 | 2-C+ | 10-C+ | 60 | 60 |
| P032 | 130 | 2.3 | 1.9 | 2-C+ | 10-C+ | 60 | 60 |
| P033 | 130 | 1.1 | 3.1 | 2-C+ | 10-C+ | 60 | 60 |
| P034 | 130 | 2 | 1.7 | 3-C+ | 10-C+ | 60 | 60 |
| P035 | 130 | 1.6 | 1.3 | 2-3-C+ | 10-C+ | 60 | 60 |
| P037 | 130 | 3.3 | 3.2 | 2-C+ | 10-C+ | 60 | 60 |

**Supp. Table 3:** Effect of Bilateral Stimulation of the Subthalamic Nucleus on Gait Characteristics: PwPD compared to Healthy Controls.

| Characteristic | PwPD Pre DBS vs Healthy Controls |  | PwPD Post DBS vs Healthy Controls |  |
| --- | --- | --- | --- | --- |
| Overall |  |  |  |  |
| Asymmetry |  |  |  |  |
| Step length | 0.56 [0.06,1.03] ‡ |  | 0.55 [0.05, 1.03] ‡ |  |
| Step time | 0.43 [-0.05, 0.90] |  | 0.72 [0.23, 1.20] ‡ |  |
| Swing time | 0.32 [-0.14, 0.76] |  | 0.47 [0.01, 0.88] |  |
| Stance time | 0.30 [-0.18, 0.74] |  | 0.45 [-0.02, 0.85] |  |
| Dyscoordination |  |  |  |  |
| PCI | 0.81 [0.3, 1.26] ‡ |  | 1.02 [0.52, 1.46] ‡ |  |
| Other gait measures |  |  |  |  |
| Walking Speed | -0.25 [-0.73, 0.22] |  | -0.13 [-0.62, 0.33] |  |
| Clinical Subtypes |  |  |  |  |
|  | PwPD Pre DBS |  | PwPD Post DBS |  |
|  | TD vs Healthy Controls | PIGD vs Healthy Controls | TD vs Healthy Controls | PIGD vs Healthy Controls |
| Asymmetry |  |  |  |  |
| Step length | - 0.50 [-1.43, 0.23] | -0.67 [-1.35, -0.06] ‡ | -0.50 [-1.2, 0.06] | -0.60 [-1.3, 0.01] ‡ |
| Step time | - 0.16 [-0.93, 0.43] | -0.62 [-1.23, -0.03] ‡ | -0.26 [-1.07, 0.35] | -1.1 [-1.8, -0.42] ‡ |
| Swing time | -0.18 [-1.35, 0.49] | -0.47 [-1.06, 0.11] | -0.31 [-1.27, 0.27] | -0.63 [-1.2, -0.03] ‡ |
| Stance time | -0.17 [-1.32, 0.48] | -0.43 [-1.02, 0.17] | 0.17 [-1.07, 0.37] | -0.66 [-1.22, -0.03] ‡ |
| Dyscoordination |  |  |  |  |
| PCI | -0.46 [-1.16, 0.10] | -1.08 [-1.64, -0.42] ‡ | -0.57 [-1.34, -0.01] | -1.45 [-2.14, -0.75] ‡ |
| Other gait measures |  |  |  |  |
| Walking Speed | -0.13 [-0.75, 0.54] | 0.52 [-0.19, 1.07] | -0.10 [-0.68, 0.64] | 0.28 [-0.34, 0.87] |

Values expressed as mean (standard deviation)
Changes are provided as hedges' g effect sizes [confidence intervals].
\* $p < 0.05$ ; The $p$ values of the two-sided permutation t-test
Abbreviations: PCI - Phase Coordination Index; PwPD - Patients with Parkinson's disease; DBS - Deep Brain Stimulation; TD - Tremor Dominant; PIGD - Postural Instability and Gait Disorder

## References

[1] S. Grillner and A. El Manira, “Current Principles of Motor Control, with Special Reference to Vertebrate Locomotion,” Physiol Rev, vol. 100, 2020.

[2] K. Takakusaki, “Functional Neuroanatomy for Posture and Gait Control,” (in English), Journal of Movement Disorders, vol. 10, 2017.

[3] N. Georgiou, R. Iansek, J. L. Bradshaw, J. G. Phillips, J. B. Mattingley, and J. A. Bradshaw, “An Evaluation of the Role of Internal Cues in the Pathogenesis of Parkinsonian Hypokinesia,” Brain, vol. 116 (Pt 6), 1993.

[4] H. Sadeghi, P. Allard, F. Prince, and H. Labelle, “Symmetry and Limb Dominance in Able-Bodied Gait: A Review,” Gait Posture, vol. 12, 2000.

[5] R. D. Gregg, Y. Y. Dhaher, A. Degani, and K. M. Lynch, “On the Mechanics of Functional Asymmetry in Bipedal Walking,” IEEE Trans Biomed Eng, vol. 59, 2012.

[6] G. Yogev, M. Plotnik, C. Peretz, N. Giladi, and J. M. Hausdorff, “Gait Asymmetry in Patients with Parkinson’s Disease and Elderly Fallers: When Does the Bilateral Coordination of Gait Require Attention,” (in English), Experimental Brain Research, vol. 177, 2007.

[7] M. D. Humphries, J. A. Obeso, and J. K. Dreyer, “Insights into Parkinson’s Disease from Computational Models of the Basal Ganglia,” (in English), Journal of Neurology Neurosurgery and Psychiatry, vol. 89, 2018.

[8] M. Plotnik, R. P. Bartsch, A. Zeev, N. Giladi, and J. M. Hausdorff, “Effects of Walking Speed on Asymmetry and Bilateral Coordination of Gait,” (in English), Gait & Posture, vol. 38, 2013.

[9] A. Fasano, C. Schlenstedt, J. Herzog, M. Plotnik, F. E. M. Rose, J. Volkmann et al., “Split-Belt Locomotion in Parkinson’s Disease Links Asymmetry, Dyscoordination and Sequence Effect,” Gait Posture, vol. 48, 2016.

[10] M. Plotnik, N. Giladi, and J. M. Hausdorff, “Bilateral Coordination of Walking and Freezing of Gait in Parkinson’s Disease,” (in English), European Journal of Neuroscience, vol. 27, 2008.

[11] M. Plotnik, N. Giladi, Y. Balash, C. Peretz, and J. M. Hausdorff, “Is Freezing of Gait in Parkinson’s Disease Related to Asymmetric Motor Function?,” (in English), Annals of Neurology, vol. 57, 2005.

[12] B. R. Bloem, J. A. Hausdorff, J. E. Visser, and N. Giladi, “Falls and Freezing of Gait in Parkinson’s Disease: A Review of Two Interconnected, Episodic Phenomena,” (in English), Movement Disorders, vol. 19, 2004.

[13] E. L. Johnsen, P. H. Mogensen, N. A. Sunde, and K. Ostergaard, “Improved Asymmetry of Gait in Parkinson’s Disease with Dbs: Gait and Postural Instability in Parkinson’s Disease Treated with Bilateral Deep Brain Stimulation in the Subthalamic Nucleus,” (in English), Movement Disorders, vol. 24, 2009.

[14] C. Miller-Patterson, R. Buesa, N. McLaughlin, R. Jones, U. Akbar, and J. H. Friedmann, “Motor Asymmetry over Time in Parkinson’s Disease,” (in English), Journal of the Neurological Sciences, vol. 393, 2018.

[15] B. W. Fling, C. Curtze, and F. B. Horak, “Gait Asymmetry in People with Parkinsons Disease Is Linked to Reduced Integrity of Callosal Sensorimotor Regions,” (in English), Frontiers in Neurology, vol. 9, 2018.

[16] P. Krack, J. Volkmann, G. Tinkhauser, and G. Deuschl, “Deep Brain Stimulation in Movement Disorders: From Experimental Surgery to Evidence-Based Therapy,” Mov Disord, 2019.

[17] A. Collomb-Clerc and M. L. Welter, “Effects of Deep Brain Stimulation on Balance and Gait in Patients with Parkinson’s Disease: A Systematic Neurophysiological Review,” Neurophysiol Clin, vol. 45, 2015.

[18] G. Cossu and M. Pau, “Subthalamic Nucleus Stimulation and Gait in Parkinson’s Disease: A Not Always Fruitful Relationship,” (in English), Gait & Posture, vol. 52, 2017.

[19] A. Castrioto, A. M. Lozano, Y. Y. Poon, A. E. Lang, M. Fallis, and E. Moro, “Ten-Year Outcome of Subthalamic Stimulation in Parkinson Disease: A Blinded Evaluation,” Arch Neurol, vol. 68, 2011.

[20] C. Schlenstedt, A. Shalash, M. Muthuraman, D. Falk, K. Witt, and G. Deuschl, “Effect of High-Frequency Subthalamic Neurostimulation on Gait and Freezing of Gait in Parkinson’s Disease: A Systematic Review and Meta-Analysis,” Eur J Neurol, vol. 24, 2017.

[21] I. Cebi, M. Scholten, A. Gharabaghi, and D. Weiss, “Clinical and Kinematic Correlates of Favorable Gait Outcomes from Subthalamic Stimulation,” Front Neurol, vol. 11, 2020.

[22] E. Lai, M. Bryant, P. Luo, K. Follett, M. Stern, D. Reda et al., “Risk of Falls in Parkinson’s Disease after Deep Brain Stimulation,” (in English), Neurology, vol. 80, 2013.

[23] A. Fasano, J. Herzog, E. Seifert, H. Stolze, D. Falk, R. Reese et al., “Modulation of Gait Coordination by Subthalamic Stimulation Improves Freezing of Gait,” (in English), Movement Disorders, vol. 26, 2011.

[24] G. T. Stebbins, C. G. Goetz, D. J. Burn, J. Jankovic, T. K. Khoo, and B. C. Tilley, “How to Identify Tremor Dominant and Postural Instability/Gait Difficulty Groups with the Movement Disorder Society Unified Parkinson’s Disease Rating Scale: Comparison with the Unified Parkinson’s Disease Rating Scale,” (in English), Movement Disorders, vol. 28, 2013.

[25] N. Konig, N. B. Singh, J. von Beckerath, L. Janke, and W. R. Taylor, “Is Gait Variability Reliable? An Assessment of Spatio-Temporal Parameters of Gait Variability During Continuous Overground Walking,” (in English), Gait & Posture, vol. 39, 2014.

[26] M. Plotnik, N. Giladi, and J. M. Hausdorff, “A New Measure for Quantifying the Bilateral Coordination of Human Gait: Effects of Aging and Parkinson’s Disease,” Exp Brain Res, vol. 181, 2007.

[27] J. Ho, T. Tumkaya, S. Aryal, H. Choi, and A. Claridge-Chang, “Moving Beyond P Values: Data Analysis with Estimation Graphics,” (in English), Nature Methods, vol. 16, 2019.

[28] J. W. Cohen, Statistical Power Analysis for the Behavioral Sciences, Second Edition ed. 1988, pp. XXI, 567 Seiten.

[29] M. J. Forjaz, A. Ayala, C. M. Testa, P. G. Bain, R. Elble, D. Haubenberger et al., “Proposing a Parkinson’s Disease-Specific Tremor Scale from the Mds-Updrs,” Mov Disord, vol. 30, 2015.

[30] A. van der Hoorn, H. Burger, K. L. Leenders, and B. M. de Jong, “Handedness Correlates with the Dominant Parkinson Side: A Systematic Review and Meta-Analysis,” Mov Disord, vol. 27, 2012.

[31] A. Fasano, C. C. Aquino, J. K. Krauss, C. R. Honey, and B. R. Bloem, “Axial Disability and Deep Brain Stimulation in Patients with Parkinson Disease,” Nat Rev Neurol, vol. 11, 2015.

[32] A. Mirelman, P. Bonato, R. Camicioli, T. D. Ellis, N. Giladi, J. L. Hamilton et al., “Gait Impairments in Parkinson’s Disease,” Lancet Neurol, vol. 18, 2019.

[33] S. Del Din, M. Elshehabi, B. Galna, M. A. Hobert, E. Warmerdam, U. Suenkel et al., “Gait Analysis with Wearables Predicts Conversion to Parkinson Disease,” (in English), Annals of Neurology, 2019.

[34] A. A. Moustafa, S. Chakravarthy, J. R. Phillips, A. Gupta, S. Keri, B. Polner et al., “Motor Symptoms in Parkinson’s Disease: A Unified Framework,” (in English), Neuroscience and Biobehavioral Reviews, vol. 68, 2016.

[35] K. J. Lizarraga, C. C. Luca, A. De Salles, A. Gorgulho, A. E. Lang, and A. Fasano, “Asymmetric Neuromodulation of Motor Circuits in Parkinson’s Disease: The Role of Subthalamic Deep Brain Stimulation,” Surg Neurol Int, vol. 8, 2017.

[36] G. Deuschl and P. Krack, “Intrepidly Studying Deep Brain Stimulation in Patients with Parkinson’s Disease,” Lancet Neurol, vol. 19, 2020.

[37] M. Katz, M. S. Luciano, K. Carlson, P. Luo, W. J. Marks, P. S. Larson et al., “Differential Effects of Deep Brain Stimulation Target on Motor Subtypes in Parkinson’s Disease,” (in English), Annals of Neurology, vol. 77, 2015.

[38] M. A. Hobert, S. Nussbaum, T. Heger, D. Berg, W. Maetzler, and S. Heinzel, “Progressive Gait Deficits in Parkinson’s Disease: A Wearable-Based Biannual 5-Year Prospective Study,” Front Aging Neurosci, vol. 11, 2019.

[39] L. A. Malone, A. J. Bastian, and G. Torres-Oviedo, “How Does the Motor System Correct for Errors in Time and Space During Locomotor Adaptation?,” (in English), Journal of Neurophysiology, vol. 108, 2012.

